# Unbiased serology reveals autoimmunity and HIV antibody signatures in HIV CNS Escape

**DOI:** 10.1101/2022.12.13.22283439

**Authors:** I.A. Hawes, B.D. Alvarenga, W. Browne, A. Wapniarski, R. Dandekar, C.M. Bartley, G.M. Sowa, J.L. DeRisi, P. Cinque, A.N. Dravid, S.J. Pleasure, M. Gisslen, R.W. Price, M.R. Wilson

## Abstract

**Background:** Antiretroviral therapy (ART) suppresses plasma and cerebrospinal fluid (CSF) HIV replication with occasional asymptomatic episodes of detectable HIV RNA known as asymptomatic (AS) escape. Neurosymptomatic (NS) CSF escape is a rare exception in which CNS HIV replication occurs in the setting of neurologic impairment. The origins of NS escape are not fully understood.

**Methods:** Using a large cohort of PLWH (n=111), including elite controllers (n=4), viral controllers (n=4), ART untreated subjects (n=18), HIV-associated dementia (n=4), ART suppressed (n=16), AS escape (n=19), NS escape (n=35), secondary escape (n=5) subjects, and HIV-negative controls (n=6), we investigated immunoreactivity to self-antigens in the CSF of NS escape by employing neuroanatomic CSF immunostaining and massively multiplexed self-antigen serology (PhIP-Seq). Additionally, we utilized pan-viral serology (VirScan) to deeply profile the CSF anti-viral antibody response and metagenomic next-generation sequencing (mNGS) for pathogen detection.

**Results:** We detected Epstein-Barr virus (EBV) DNA more frequently in the CSF of NS escape subjects than in AS escape controls. Based on immunostaining and PhIP-Seq, there was evidence for increased immunoreactivity against self-antigens in NS escape CSF. Finally, VirScan revealed several immunodominant epitopes that map to the HIV env and gag proteins in the CSF of PLWH.

**Discussion:** We deployed agnostic tools to study whether there was evidence for a neuroinvasive co-infection and/or autoimmunity in HIV escape syndromes. We more frequently detected EBV DNA and immunoreactivity to self-antigens in NS escape. Whether these additional inflammatory markers are byproducts of an HIV-driven inflammatory process or whether they independently contribute to the neuropathogenesis of NS escape will require further study.

## Introduction

Infection of the central nervous system (CNS) begins early and continues throughout the course of systemic HIV-1 infection in the absence of anti-retroviral therapy (ART)^1^. In addition to its impact on systemic infection, ART also inhibits viral replication in the CNS, effectively eliminating detectable HIV from the cerebrospinal fluid (CSF)^2,3^ and is effective in preventing the most severe direct CNS complication, HIV-associated dementia (HAD)^4^. While this CNS therapeutic success is the rule, there are unusual exceptions in which systemic replication is suppressed but remains at detectable levels in CSF. This is referred to as HIV CSF escape which is subdivided into three types: asymptomatic CSF escape (AS escape), neurosymptomatic CSF escape (NS escape) and secondary CSF escape^5^.

Each of these three CSF escape syndromes is of pathobiological interest, but only NS escape has a clear, immediate clinical impact. Secondary CSF escape develops in the context of another infection or inflammatory condition within the CNS^5^. Importantly, when the other (secondary) infection resolves, so does the local HIV replication^6^. AS escape has primarily been detected in cohort studies in which volunteers undergo lumbar puncture (LP) as part of natural history studies of HIV infection, including treated infection^7,8,9^. In contrast to AS escape, NS escape directly causes CNS injury. It presents with diverse neurological symptoms and signs, at times severe^3,10^. CSF HIV RNA levels are usually higher than in AS escape, and neuroimaging is frequently abnormal^10^. The prevailing concept is that HIV encephalitis develops in these individuals because of some combination of i) reduced treatment adherence and ii) local drug resistance^7^.

Since HIV CSF escape is rare, the question has arisen whether other, yet-undefined factors may predispose to or provoke NS escape, including the presence of other pathogens such as Epstein-Barr virus (EBV) or cytomegalovirus (CMV)^5,11^ or a systemic or CNS autoimmune process, with its own potential for injury. In order to investigate these possibilities, we interrogated CSF samples from an array of people living with HIV (PLWH), including those with NS escape, AS escape, and secondary CSF escape. Metagenomic next-generation sequencing (mNGS)^12^ and pan-viral autoantibody profiling using programmable phage display (VirScan)^13,14^ were used to identify other infectious agents in an unbiased manner. We looked broadly for autoantigens using rodent brain tissue-based immunofluorescence and pan-human proteome programmable phage display (PhIP-Seq)^15,16^.

## MATERIALS AND METHODS

### Subject enrollment

The samples used in this study were collected between 2000 and 2018 in the context of HIV research studies at two medical centers: Sahlgrenska University Hospital, Gothenburg, Sweden and Zuckerberg San Francisco General Hospital, University of California San Francisco, San Francisco, CA, USA^17^. The initiating focus of this study was on NS escape (n=35), but we included a wide array of comparison groups (Table 1 and Figure 1A).

**Table 1:**
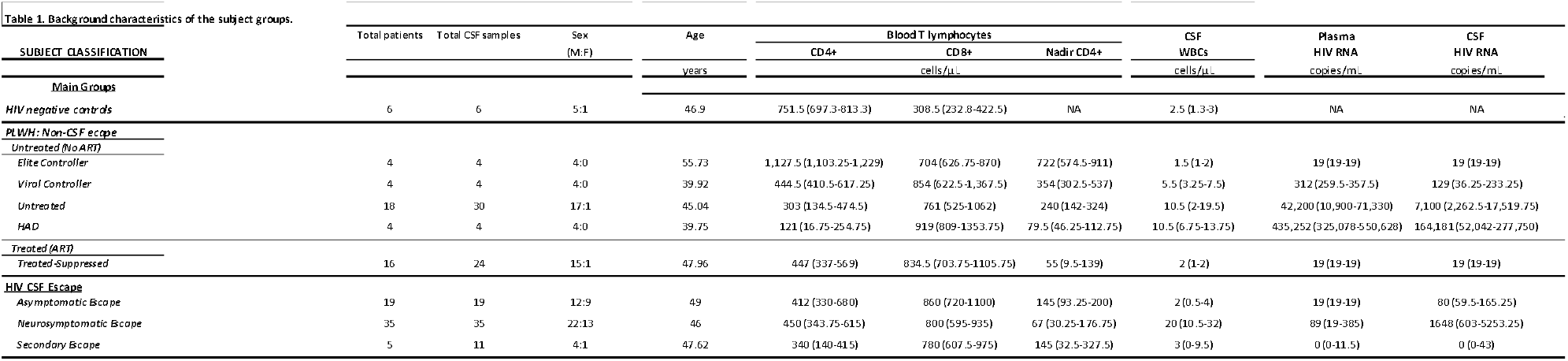
Background characteristics of subject groups.

**Fig 1.**
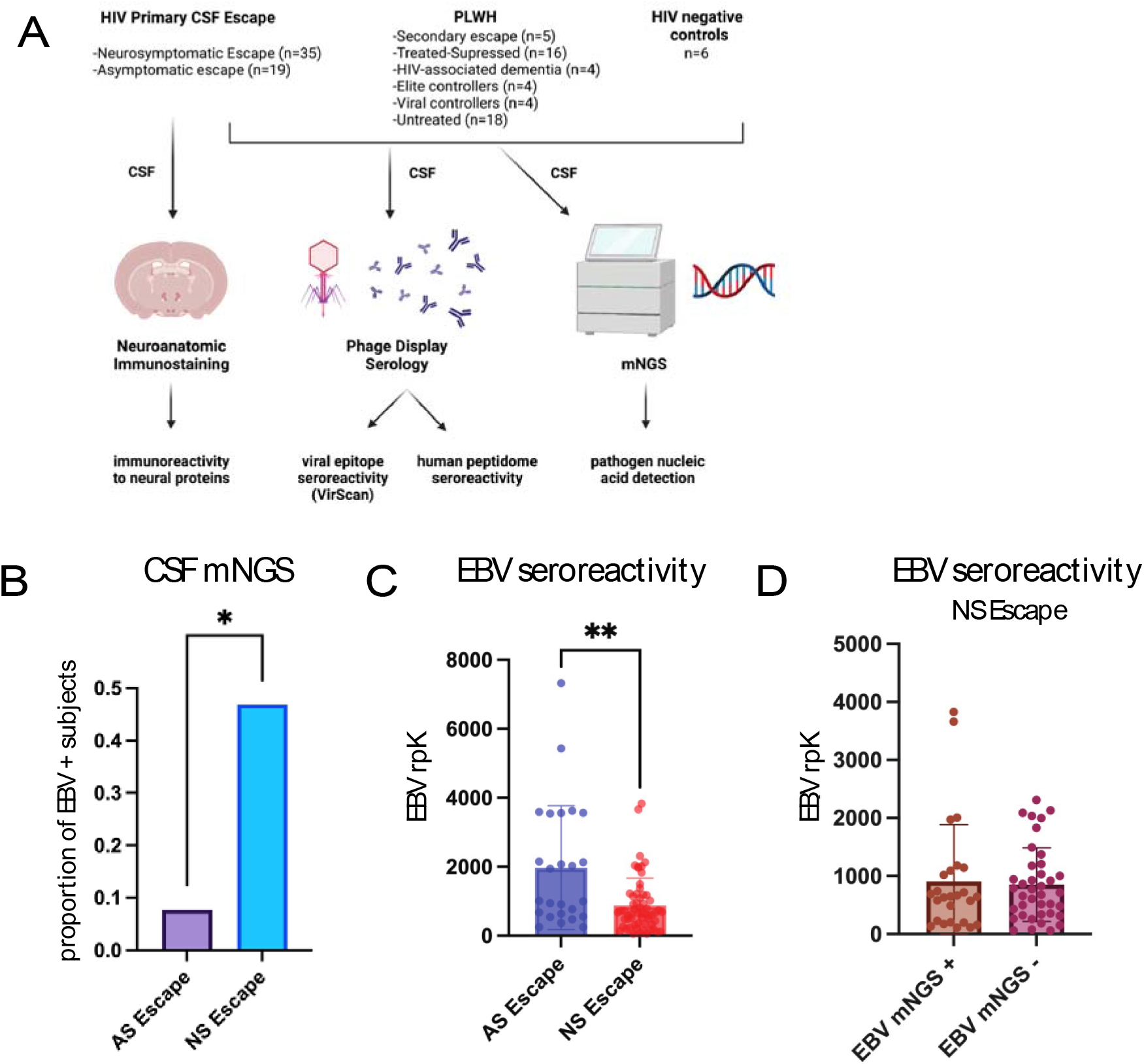
CSF mNGS detects a higher burden of EBV in NS Escape subjects. A.) Schematic of the study design. PLWH with diverse neurologic phenotypes, including NS and AS escape, underwent a lumbar puncture. CSF autoantibody and anti-viral antibody profiling was done using phage display assays (human peptidome and VirScan) and neuroanatomic staining on rodent brain tissue.Unbiased metagenomicsequencing was done on DNA extracted from CSF. B.) Proportion of subjects with Epstein-Barr virus (EBV) detected by CSF mNGS in NS escape (n=32) and AS escape (n=13) subjects. C.) Quantification of EBV seroreactivity by VirScan in AS and NS escape subjects (in replicate). Results are reported in reads per 100,000 (rpK). D.) Quantification of EBV seroreactivity by VirScan in NS escape subjects who had EBV detected in their CSF by mNGS versus those where EBV was not detected by mNGS. Statistical analyses were done using a Mann-Whitney U test in panels C and D. Statistical analysis for panel B was done using a chi-square test. Data represents mean rpK +/- SEM.

### mNGS for pathogen detection

DNA sequencing libraries were prepared using a previously described protocol optimized and adapted for miniaturization^18^, and libraries were sequenced on a NovaSeq 6000 machine (Illumina) to generate 150 nucleotide (nt), paired-end reads. Candidate DNA virus pathogens were identified from raw mNGS sequencing reads using CZID v3.2, a cloud-based, open-source bioinformatics platform designed for detection of microbes from mNGS data^19^.

### VirScan (pan-viral) and peptidome (pan-human proteome) serologic assays

PhIP-Seq assays to identify anti-viral antibodies (i.e., VirScan) and autoantibodies were performed as previously described in technical replicate^14,20^. Briefly, phage display libraries were amplified in *E. coli* and incubated overnight with 2uL of subject CSF or serum and underwent two rounds of immunoprecipitation. Barcoded phage DNA were then sequenced on a NovaSeq 6000 (Illumina).

### Bioinformatic Analysis of Phage Data

For the VirScan and human peptidome phage analyses, data were analyzed using previously described methods^14,20^ (Supplemental Methods). Briefly, peptide sequences were aligned to the reference library, and counts were normalized by creating a ratio of specific peptide reads to the number of sequencing reads and multiplying by 100,000 to create a measure of reads per hundred thousand (rpK). rpKs for individual peptides in each subject were filtered using a cutoff fold-change (FC) of greater than 10 above the mean background rpK generated from null IPs. Peptide position mapping was performed by aligning peptide sequences in R Studio using the msa (multiple sequence alignment) Bioconductor package^21^ and using the envelope protein and gag/pol polyprotein portions of the HXB2 HIV clade B sequence (GenBank K03455.1) as reference sequences.

### Anatomic Mouse Brain Staining

Mouse brain sections fixed overnight in 4% paraformaldehyde were immunostained with CSF at 1:25 dilution were prepared as previously described^20^ (Supplemental Methods). CSF immunostaining was assessed by two separate scientists (SJP and WB) experienced in immunohistochemistry analysis in a blinded manner.

### Statistical Methods

All data comparisons between two subsets of subjects were made using the Mann-Whitney test or Chi-square test, and exact n and error bars are provided and defined in figure legends. All comparisons involving two or more subsets of subjects were performed with the ANOVA test.

## Data Availability

Anonymized data not published within this article will be made available by request from any qualified investigator.

## RESULTS

### Features of the study cohort

This study included PLWH (n=111) including elite controllers (n=4), viral controllers (n=4), ART untreated subjects (n=18), HIV-associated dementia (n=4), ART suppressed (n=16), asymptomatic CSF escape (n=19), neurosymptomatic escape (n=35), and secondary escape (n=5) subjects as well as HIV-negative controls (n=6). The salient HIV-related background characteristics of the study cohort are summarized in **Table 1**. The HIV CSF escape groups adhered to recent definitions^22^ and the samples were obtained in the context of clinical presentation (NS escape and HAD). Subjects enrolled as part of cohort studies (all other subjects) and have been described previously^17,23,24^.

### CSF mNGS detects a higher burden of EBV in NS escape subjects

We sequenced 45 CSF samples from AS and NS escape subjects using DNA mNGS with a median sequencing depth of 41,163,352 reads per sample (IQR 16,384,312-133,365,998 reads per sample). There was a statistically significant increase in EBV detection in the of NS escape subjects (n=13 out of 32) when compared to AS escape subjects (n=1 out of 13) (Chi-square test, p<0.05) (Figure 1b). In addition to EBV detection (0.1-0.8 reads per million (rpM)), we detected CMV and human herpes virus-8 (HHV-8) in one NS escape subject and HHV-6 in another NS escape subject (Supp Table 1). Aside from human herpes viruses, no other neuroinvasive viruses were detected in any of the 45 subjects.

Using our VirScan platform, AS escape subjects had a significantly higher average enrichment of EBV-specific peptides relative to NS escape subjects (Figure 1c). Relative to AS escape subjects, the CSF of NS escape subjects did not enrich viral peptides in non-retroviridae/non-herpesviridae viral families (Supp. Figure 1b). There was no difference in enrichment for EBV-specific peptides in NS escape subjects for whom EBV nucleic acid was detected in the CSF versus those in whom EBV was not detected (Figure 1d).

### The anti-HIV antibody repertoire suggests compartmentalization of the anti-HIV immune response

VirScan detected enrichment of HIV-specific antibodies in the CSF and serum of all study subjects with paired CSF and serum available relative to non-HIV infected controls. Anti-HIV antibodies on average were increased as a percentage of total viral antibodies in the CSF (n=85) (median 42,336 rpK, IQR 31,751-53,245 rpK) versus serum (n=78) (median 8335 rpK, IQR 5636-11687 rpK) (p< 0.0001) across our entire cohort (Figure 2a).

**Fig 2.**
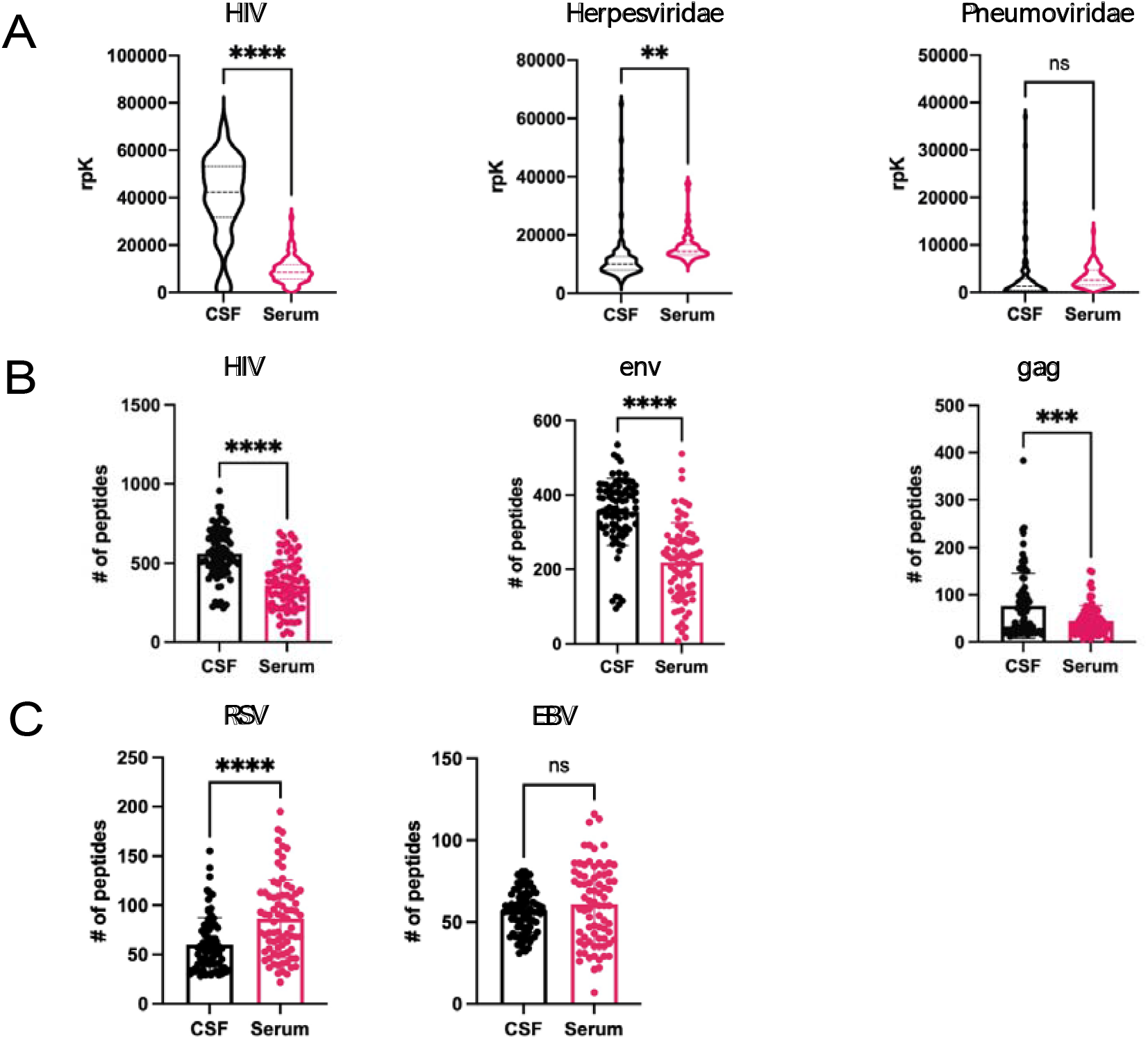
The anti-HIV antibody repertoire suggests CNS compartmentalization of the anti-HIV immune response not reflected in other viral families. Pan-viral serology (VirScan) was performed on CSF samples from PLWHIV with diverse neurologic phenotypes and treatment status. Results are reported in reads per 100,000 (rpK). A.) Violin plots of reads mapping to HIV, Herpesviridae, and Pneumoviridae respecitvely in matched CSF and serum B.) Bar graphs of the number of unique peptides mapping to the HIV genome, env protein, and gag protein respectively that were detected in matched CSF and serum C.) Bar graphs of the number of unique peptides mapping t o respiratory syncytial virus (RSV) and Epstein-Barr virus (EBV) respectively that were detected in matched CSF and serum. All statistical analyses were done using a Mann-Whitney U test. Data represents mean rpK+/- SEM.

To examine whether this effect was HIV-specific, we examined whether peptides derived from Herpesviridae and Pneumoviridae were similarly enriched using CSF versus serum in the PLWH cohort. Unlike HIV, reactivity to Herpesviridae and Pneumonviridae species were not increased as a percentage of total viral antibodies in the CSF relative to serum (Figure 2a).

Next, in order to assess the degree of compartmentalization of the anti-HIV antibody repertoire, the diversity of HIV peptides enriched using CSF and serum was examined. CSF enriched a greater number of unique HIV peptides than serum samples (CSF median 556, IQR 460.5-679, serum median 350.5, IQR 227-474.8) (p< 0.0001) (Figure 2b). To examine whether this effect was HIV-specific, two common non-retroviruses, respiratory syncytial virus (RSV) and EBV were examined. Unlike HIV, neither yielded a differential enrichment of peptides in CSF compared to serum. (Figure 2c).

### VirScan shows a conserved immunodominant epitope signature in the HIV envelope protein

In HIV, the humoral immune response is primarily directed against the HIV envelope protein and the gag/pol polyprotein^25^. In our cohort, VirScan identified two conserved immunodominant envelope protein epitopes starting at amino acid (AA) positions 289 and 577 (Figure 3a). The AA 289 epitope maps to the V3 loop of the envelope protein, which is an important determinant of X4/R5 tropism of viral quasi-species^26^. The epitope at AA 577 maps to the C-terminal heptad repeat 2 region (CH2 terminal), which plays an important role in CD4 receptor binding^27^. We found that CSF immunoglobulins enriched HIV envelope peptides from AS escape subjects (n=19) to a greater degree than NS escape subjects (n=35) (AS escape median 64,567 rpK IQR 56,306-69,994 rpK vs NS escape median 35,734 rpk IQR 22,536 -47,053 rpK) (p<0.001) (Figure 3b). This difference is largely driven by an enrichment of the AA 577 CH2 epitope in AS escape subjects.

**Fig 3.**
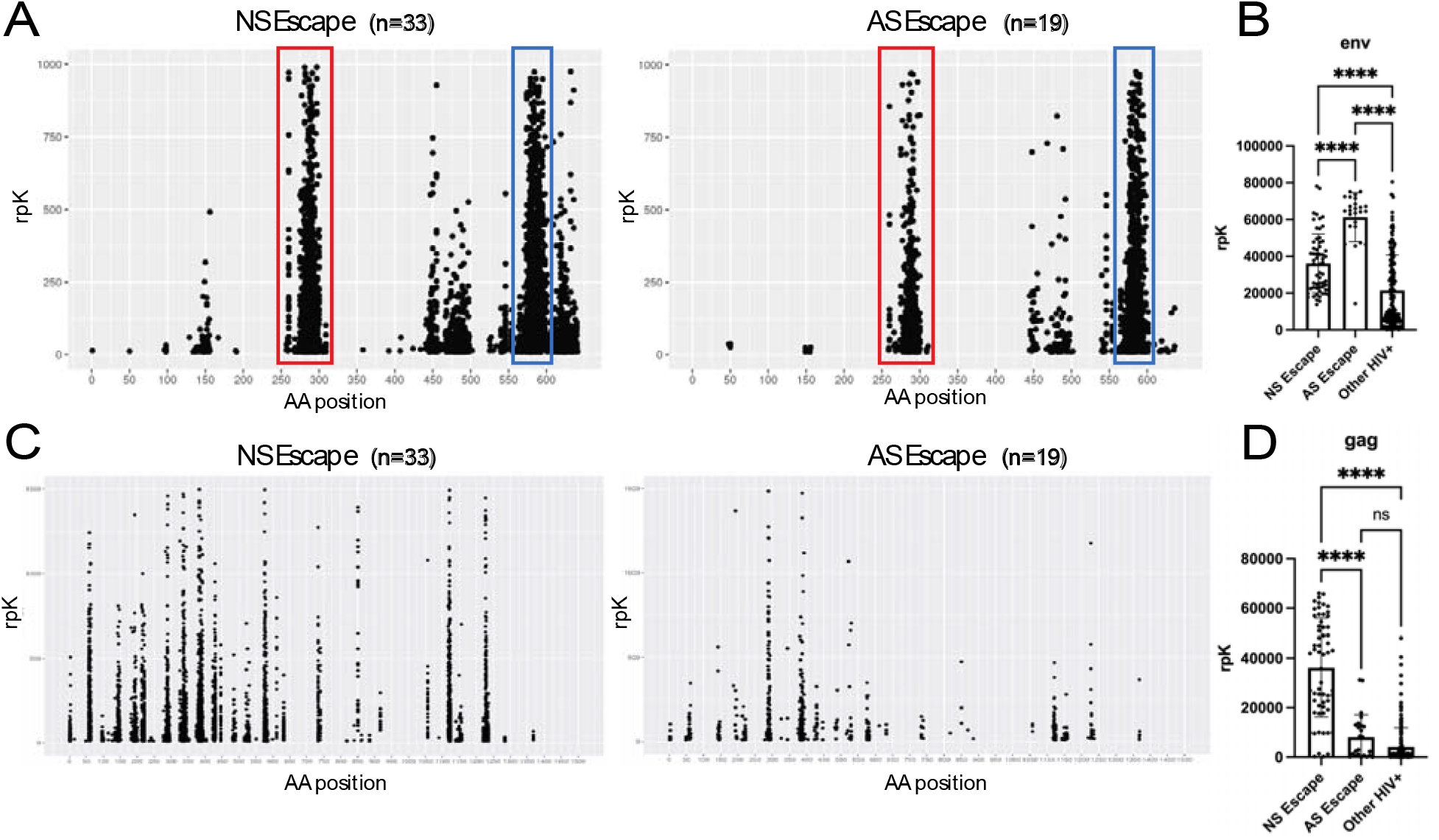
Deep epitope profling of HIV antigenic specifcity in the CSF reveals differential epitope targeting between AS and NS escape subjects. A.) CSF antibody epitope maps of the HIV env protein from AS and NS escape subjects. Each point on the graph represents the targeting of a single peptide from a single subject. Epitopes of note at AA position 289 (red) and AA position 577 (blue) are highlighted B.) Quantifcation of the summed rpK of peptides mapping t o the env protein enriched by CSF samples from subjects with AS and NS escape, as well as other PLWH. C.) CSF antibody epitope maps of the HIV gag protein from AS and NS escape subjects.Each point on the graph represents the targeting of a single peptide from a single subject D.) Quantifcation of the rpK of peptides mapping to the gag protein enriched by CSF from subjects with AS and NS escape, as well as other PLWH. All statistical analyses were done using a one way ANOVA test. Data represents mean rpK +/- SEM.

### VirScan shows increased antibody reactivity to the gag protein in subjects with NS escape compared to AS escape

In contrast to our envelope protein findings, epitope mapping of the gag protein across PLWH shows that antibody specificity was much more diffuse (Figure 3c). Enrichment of gag-derived peptides did not correlate with treatment status or most neurological phenotypes. However, we found that there was a significant enrichment of HIV gag peptides in NS escape (median rpK 41,770, IQR 20,493-54051 rpK) compared to AS escape cases (median rpK 2,953, IQR 673-12848 rpK) (p<0.001) (Figure 3d). The difference in HIV gag protein enrichment in NS escape subjects was largely driven by peptides starting at AA position 433, mapping to the gag spacer peptide 2 (p1) domain, which is a recently discovered, protective T-cell epitope^28^.

### CSF from some NS escape subjects demonstrate immunoreactivity to rodent brain tissue

Of the 32 NS escape cases whose CSF was immunostained, 10 were classified as positive (Figure 4a). In contrast, only one case out of 19 in the AS escape group was classified as positive. There were some commonalities in the brain regions that demonstrated staining (Fig 4b-c).

**Fig 5.**
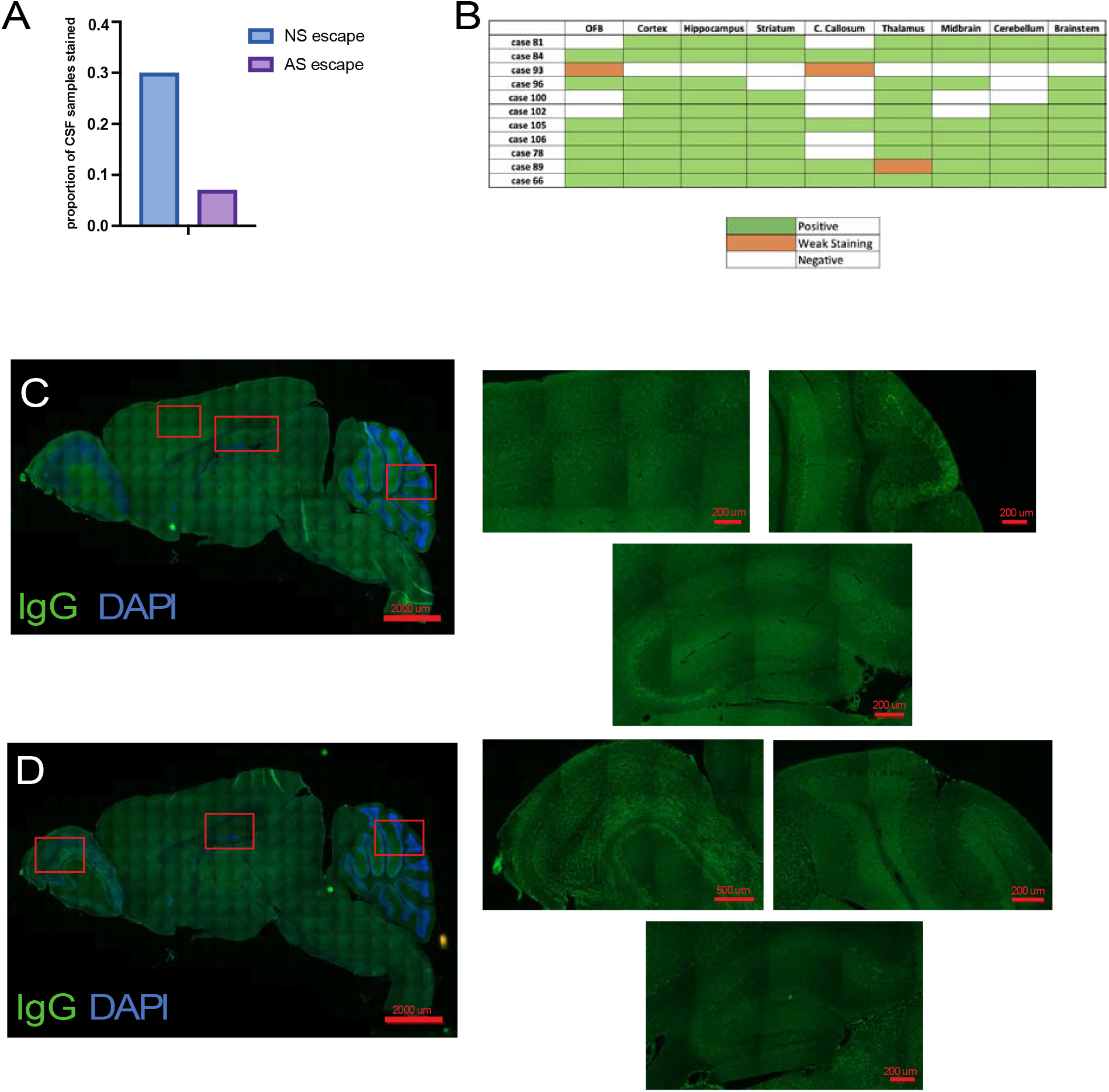
Increased neuroanatomic immunoreactivity in NS escape CSF suggests increased prevalence of anti-neural antibodies. A.) Quantification of proportion of NS escape (n=33) and AS escape (n=13) subject CSF that had a positive staining pattern on the anatomic staining platform. B.) Matrix indicating brain regions with anatomic immunoreactivity of NS and AS escape CSF C.) Whole brain AxioScan image of a sagittal slice of brain tissue stained at a 1:25 dilution with CSF (green) from NS escape subject case 105 and the nuclear stain DAPI (blue). Additional high-resolutionrepresentative images are shown from cortex (top left), cerebellum (top right) and hippocampus (bottom). D.) Whole brain AxioScan image of a sagittal slice of brain tissue stained 1:25 with patient CSF from NS escape patient case 106. Additional high-resolution representative images are shown from cortex (top left) cerebellum (top right) and hippocampus (bottom).

### NS escape subjects show increased number of candidate autoantibodies in the CSF by PhIP-Seq

NS escape subjects had significantly higher numbers of candidate CSF autoantigens (median: 39 genes enriched, IQR 23-54) than AS escape controls (median: 23 genes enriched, IQR 11.5-32) (p<0.0001) (Figure 5b).

**Fig 5.**
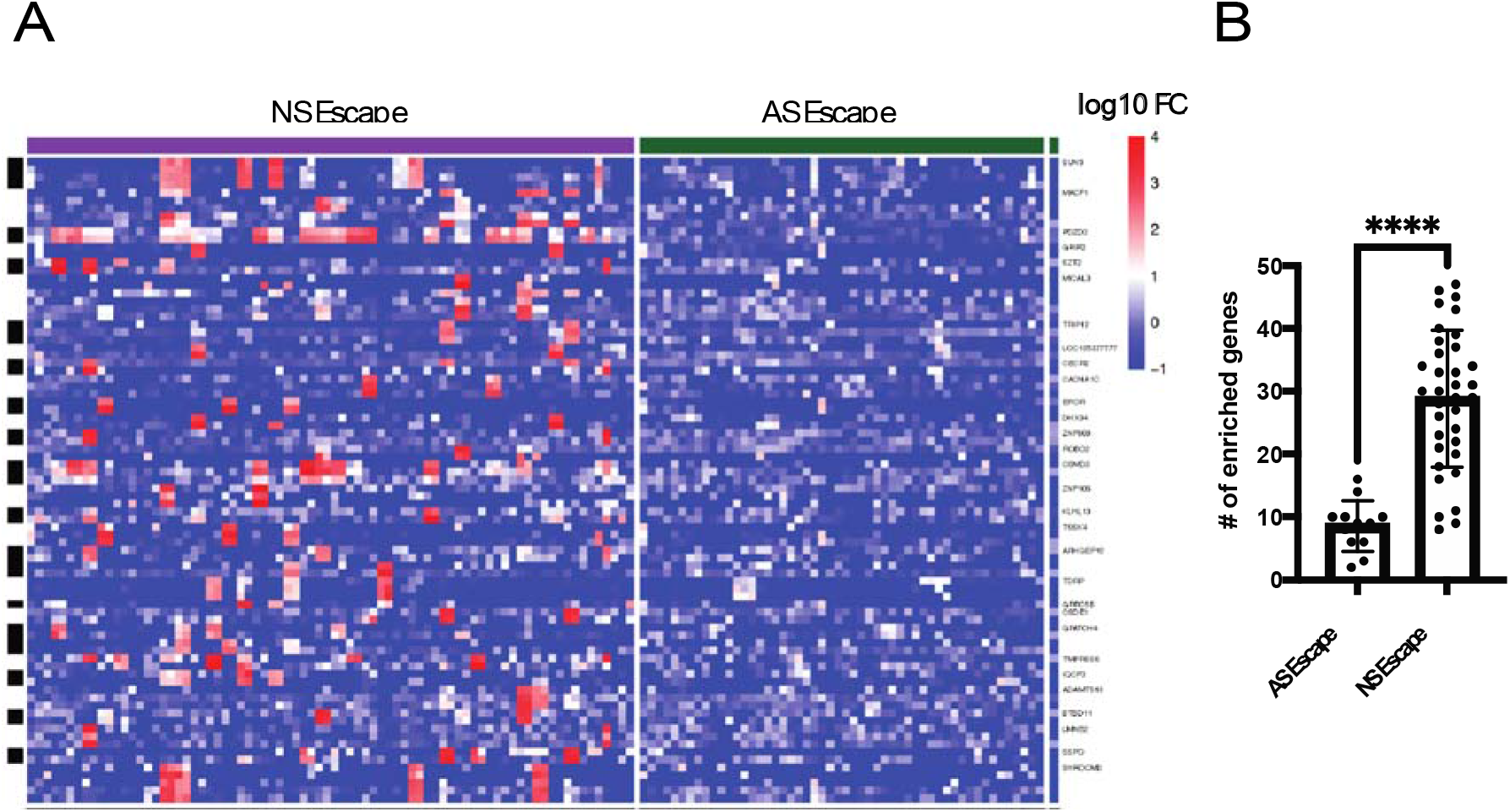
NS escape CSF shows increased autoantibody prevalence by PhIP-Seq. CSF from ASand NSescape subjects was run on a pan-human peptidome serology assay. Proteins were considered candidate autoantigens if they had at least one peptide enriched relative to a null template AG bead control and non-HIV CSF escape background (see methods) A.) Heatmap of genes in AS (n=19) and NS escape (n=33) cases that were enriched above background. Each column is a single subject sample (grouped by NS and AS escape) and each row is a single enriched peptide (grouped by gene) B.) Quantification of the number of genes in AS and NS escape cases that were enriched above background. All statistical analyses were done using a Mann-Whitney U test. Data represents mean rpK +/- SEM.

While we did not find evidence of a shared or “public” autoantigen in NS escape subjects, peptides from CNS associated proteins were enriched across multiple NS escape subjects, including SZT2 (n=4) with an average FC of 239.8, ROBO2 (n=2) with an average FC of 152.6, ARSA (n=2) with an average FC of 30.42, and MACF1 (n=8) with an average FC of 227.4.

## Discussion

Neurological symptoms usually abate in NS escape subjects when ART is adjusted, paralleled with decreased CSF HIV RNA concentrations^29^. This provides strong evidence for HIV-1 as the etiologic agent. However, there are HIV subjects with increased HIV replication in the CSF with no neurological symptoms (AS escape)^9^, calling into question whether the pathogenesis of NS escape can be solely explained by CNS HIV replication. Here, we deployed a number of advanced tools in PLWH with (and without) a variety of neurological complications to study whether there was evidence for a neuroinvasive co-infection and/or autoimmunity.

With regard to potential co-infections, we detected EBV in the CSF at significantly higher rates in NS escape subjects compared to AS escape subjects using DNA mNGS. This finding could reflect an increased B-cell abundance in NS escape samples. Indeed, CSF white blood cell counts are usually higher in NS compared to AS escape, which was also the case in our cohort (Table 1). This finding is also consistent with studies showing that EBV positivity in PLWH correlates with higher HIV RNA levels and CNS inflammation^11,30^. As a complement to mNGS, we utilized pan-viral antibody profiling of CSF with VirScan. We found that, paradoxically, AS escape subjects displayed increased enrichment of EBV-specific peptides compared to NS escape subjects. Since VirScan is only semi-quantitative and broadly detects both neutralizing and non-neutralizing anti-viral antibodies, it remains an open question whether this decrease in EBV peptide enrichment in NS escape subjects actually reflects a less effective humoral response to EBV.

We showed an enrichment of anti-HIV specific antibodies in CSF relative to serum, as well as a greater number of unique anti-HIV epitopes targeted by CSF antibodies relative to serum. This may simply reflect the greater diversity of humoral anti-viral responses in the periphery relative to the CNS. However, these results could also suggest a broader immune response to HIV in the CNS, potentially due to viral quasi-species that can be unique to the CNS^31^. Indeed, a recent study by Spatola et al^32^ identified antibodies in the CSF of chronically infected PLWH that were compartmentalized. They found that CSF IgG1 and IgG3 antibodies had functionally ineffective Fc-effector profiles that were not found in the plasma.

We found two immunodominant HIV envelope protein epitopes in both the CSF and in the serum that map to AA position 289 in the V3 region and AA position 577 in the CH2 terminal region. The V3 region is integrally involved in both receptor binding and the determination of viral tropism^33,34^. The CH2 terminal region plays a key role in HIV-1 entry by membrane fusion^35^. When we compared the enrichment of these envelope peptides between NS and AS escape subjects, we found that AS escape subjects more highly enriched the CH2 terminal epitope relative to NS escape subjects. In addition to these conserved envelope peptides, we also found a gag peptide at AA position 433 that is significantly enriched in NS escape subjects relative to AS escape subjects. This peptide maps to the gag spacer peptide 2 (p1) domain, which is a recently discovered, protective T-cell epitope^28^. Notably, all three of these epitopes (env AA positions 289 and 577, gag AA position 433) in the CSF of PLWH were also identified by Eshleman et al^36^ in their VirScan study profiling anti-HIV antibodies in sera from 57 PLWH. This study identified 4 antigens that were associated with disease duration, and three of those four epitopes are contained within the V3 and CH2 terminal regions we identified as well as the gag epitope that was enriched in NS escape subjects. Whether these differences in enrichment represent a functionally consequential difference in the humoral immune response between NS escape and AS escape needs further study.

HIV subjects can exhibit broad B-cell dysfunction even in the presence of ART^25^. Here, we found that the CSF of NS escape subjects demonstrated increased rodent brain immunoreactivity when compared to AS escape subjects, suggesting an increased burden of anti-neural antibodies. Additionally, PhIP-Seq identified candidate CNS-associated autoantigens, including SZT2, ROBO2, ARSA, and MACF1 across multiple NS escape subjects. Interestingly, MACF1 has been shown to regulate microtubule and actin dynamics in developing neurons, and mutations in this gene have been associated with lissencephaly^37^. These results suggest an increased burden of CNS autoimmunity in NS escape subjects when compared to their AS escape counterparts. However, it is unclear whether these differences are a cause or a consequence of NS escape, especially given that many of the candidate autoantigens are intracellular proteins, and as a result, antibodies targeting them may not be pathogenic.

This study has a number of limitations. While we identified a higher number of NS escape subjects versus AS escape subjects with EBV nucleic acid in CSF, we cannot rule out the possibility of other co-infections present in NS escape subjects. Because we performed mNGS on CSF DNA, we were insensitive to detecting neuroinvasive RNA viruses. Our VirScan assay preferentially immunoprecipitates IgG and not IgM antibodies, making the assay less sensitive to an acute humoral anti-viral response, In addition, these subjects were immunosuppressed and may have had a blunted humoral immune response. Additionally, antibodies targeting conformational or post-translationally modified antigens are not well detected by our mostly linear peptide VirScan and human peptidome assays. Furthermore, these studies were performed on CSF that in some cases was over two decades old and had undergone multiple freeze thaws, raising the potential for sample degradation. Finally, our candidate autoantigens need to be validated using orthogonal assays and assessed in prospective cohorts to determine whether they might serve as biomarkers of disease and/or contribute to disease pathogenesis.

We deployed agnostic tools in PLWH with a variety of neurological complications to assess for evidence of a neuroinvasive co-infection and/or autoimmunity that might enhance our understanding of HIV escape syndromes. In the CSF, we more frequently detected EBV DNA and immunoreactivity to self-antigens in NS escape subjects compared to AS escape subjects. Whether these additional inflammatory markers are byproducts of an HIV-driven inflammatory process associated with neurologic impairment or whether they independently contribute to the neuropathogenesis of NS escape will require further study.

## Acknowledgements

Sequencing was performed at the UCSF Center for Advanced Technology, supported by UCSF PBBR, RRP IMIA, and NIH 1S10OD028511-01 grants. We thank Thomas Ngo for his guidance on the rodent brain anatomic immunostaining platform.

## SUPPLEMENTARY MATERIALS AND METHODS

### Subject enrollment

The samples used in this study were collected between 2000 and 2018 in the context of HIV research studies at two medical centers: Sahlgrenska University Hospital, Gothenburg, Sweden and Zuckerberg San Francisco General Hospital, University of California San Francisco, San Francisco, CA, USA^17,23,24^. Samples were obtained in the context of established research approved by the local institutional review boards as previously described^17,23,24^. All participants provided informed consent and, if there was a question regarding their capacity to provide fully consent, consent was also obtained from individuals with power of attorney.

The initiating focus of this study was on NSE (n=35), but we included a wide array of comparison groups, including the following HIV-infected groups: AS escape (n=19); secondary CSF escape (n=5); untreated-viremic (n=18); untreated HAD (n=4); untreated viral controllers (n=4) and elite controllers (n=4); and treated virally suppressed (n=16). Six HIV-uninfected controls were also included (Table 1). In brief, NSE subjects presented with new or progressive CNS deficits and found to have the CSF-blood profiles defining that entity; most had imaging abnormalities. AS escape was identified in cohort studies while four secondary CSF escape subjects were identified in Gothenburg—two with herpes zoster (cases 109 & 111), one with cytomegalovirus (case 110), and one with herpes simplex virus 2 (case 108) which were previously described^17^. Of the subjects with VZV, there was one accompanied by active CNS HIV replication and one without. The background variables in Table 1 were measured in the clinical laboratories at the two participating medical centers using standard methods. Plasma and CSF viral loads were considered ‘undetectable’ below 20 HIV RNA copies per mL and assigned a default ‘floor value’ of 16 copies per mL Elite controllers had <20 copies HIV RNA per mL in plasma while viral controllers had plasma HIV RNA concentrations between 20 and 500 copies per mL.

This selection and the various subject numbers were based largely on availability and our emphasis on a broad subject sample for comparison with the NS escape group. AS escape and NS escape samples were used in all the studies in this paper, including anatomic immunostaining with CSF. All other clinical groups were analyzed on our phage platforms. Of the secondary escape subjects, two of these subjects had VZV co-infections, one accompanied by active CNS HIV replication and one without. Two other subjects, one with CMV encephalitis and one with HSV-2 encephalitis were included as well.

### VirScan (pan-viral) and peptidome (pan-human proteome) serologic assays

Phage libraries were amplified *in vivo* in *E. coli* BLT5403, tittered by plaque assay, and adjusted to a concentration of 10^9^-10^11^ pfu/mL. Previous studies using other versions of VirScan have had approximately 3,000 unique HIV peptides in their libraries^13,36^, while our VirScan library encompasses approximately 7,500 unique HIV sequences^14^ that were derived from clade-type representative sequences in the Los Alamos database. Phage libraries were aliquoted into blocked plates, mixed with 2uL of subject sample, and incubated overnight. Subject antibodies were then immunoprecipitated (IP) using protein A and G beads (Thermo-Fisher Scientific) and washed by magnetic separation. Subsequent to final wash, beads were re-suspended and used to inoculate fresh *E. coli* cultures for in vivo amplification of antibody-bound phage. After lysis was completed, an aliquot of lysate was saved (IP1) and another aliquot was mixed with an additional 2uL of subject sample overnight and immunoprecipitation was repeated (IP2) to further enrich for relevant antibody-binding phage. After IPs were completed, enrichment of phage DNA and barcoding of individual IP reactions was performed using a single PCR reaction with multiplexing primers. PCR products were subsequently pooled, and bead cleaned to remove primers and size-select the final product (SPRISelect, Beckman Coulter). Library concentration was quantified and libraries with concentrations between 1-4nM were sequenced on the NovaSeq 6000 (Illumina) using 150nt paired-end reads.

### PairSeq

We first translated and aligned raw PhIP-Seq DNA sequencing reads to a reference database of the human peptidome (v1) (derived from the NCBI human proteome, November 2015) using RAPSearch (v2.2) resulting in a matrix of peptide counts for each sample. Counts across all samples were scaled to rpK. We then used a custom R bioinformatics pipeline to identify candidate autoantigens (described below). Protein A/G beads were used as negative controls, and a commercial GFAP polyclonal antibody was used as a positive control. Cohort samples underwent two analyses (1) cohort symptomatic and asymptomatic HIV samples were compared to the non-cohort HIV positive reference group and (2) cohort symptomatic and asymptomatic HIV samples were compared to non-cohort HIV negative specimens. The analytic workflow was the same for both cohort analyses 1 and 2. The fold change (FC) for each peptide for each cohort sample was calculated by dividing the individual peptide rpK by the mean rpK for that peptide in the reference group. Only peptides with a FC ≥ 10 were considered “enriched” and passed forward for further analysis. Enriched peptides then underwent a 7 amino acid (AA) k-mer analysis. Enriched peptides that shared at least one identical 7AA sequence with at least one peptide from the same gene were kept for downstream analysis. Nonoverlapping enriched peptides were kept if they had an FC ≥ 100 and an rpK value ≥ 20. After the above steps, proteins were considered candidate autoantigens if they had at least one peptide with a FC ≥ 100 and a total protein rpK > 50 in both analysis 1 and 2. We also screened for autoantigens enriched at the cohort level by comparing the mean rpK for a given peptide within a cohort to the reference group and applying the same parameters as above. Plots for the top enriched peptides in both HIV symptomatic and HIV asymptomatic subjects were generated using R. Only peptides enriched in at least three disease samples were visualized.

### Anatomic Mouse Brain Staining

Mouse brain sections for anatomic immunostaining were prepared as previously described^20^. In this study, sections were incubated with CSF at a 1:25 dilution overnight at 4°C. Sections were washed 5x with PBS and stained with anti-human IgG (Alexafluor 488, cat# 709-545-149) as a secondary antibody and DAPI as a nuclear marker at 1:10,000. Panoramic epifluorescent images of stained mouse brain sections were captured at a 20x on a Zeiss AxioScan. Images were classified in a blinded manor by two independent reviewers (SJP and WB) by whether they had a distinct anatomical staining pattern (“positive”) or no staining (“negative”). Studies were approved the UCSF IACUC committee.

**Supplementary Fig 1.**
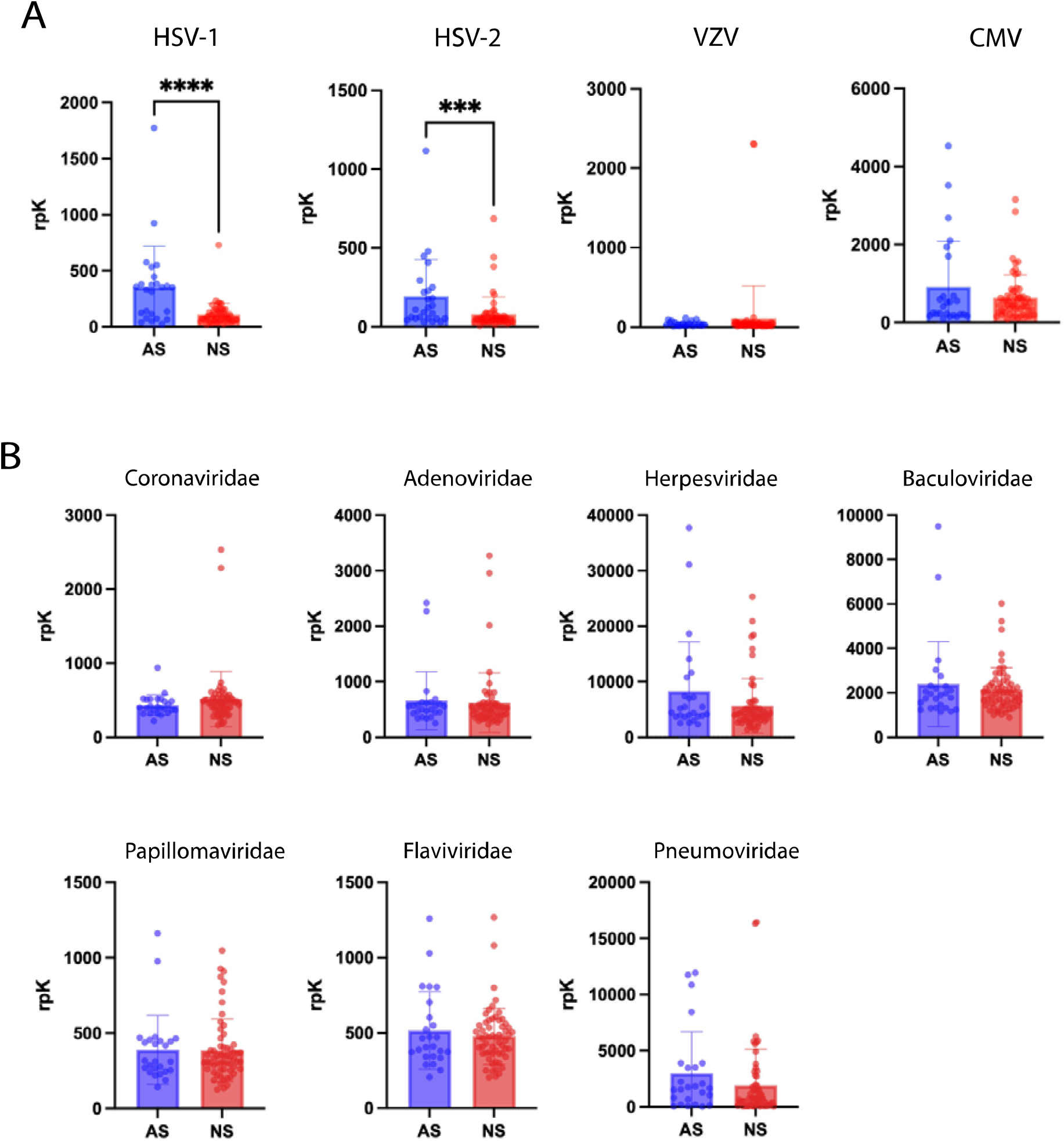
VirScan does not detect evidence of cryptic CNS co-infections in NS escape subjects. Pan-viral serology (VirScan) was performed on NS (n=33) and AS (n=19) escape CSF samples (in replicate). Results are reported in reads per 100,000 (rpK). A.) Quantification of rpK for viral species of interest in the Herpesviridae family is shown for Herpes simplex virus 1 (HSV-1),Herpes simplex virus 2 (HSV-2), Varicella zoster virus (HHV-3), Epstein-Barr Virus (HHV-4), Human Cytomegalovirus (HHV-5) B.) Quantification of rpK for viral families is shown for Coronaviridae (p=0.18), Adenoviridae (p= 0.43), Herpesviridae (p=0.12), Baculoviridae (p= 0.65), Papillomaviridae(p=0.91), Flaviviridae (p= 0.95), and Pneumoviridae (p=0.l). All statistical analyses were done using a Mann-Whitney U test. Data represents mean rpK +/- SEM.

**Supplementary Table 1:**
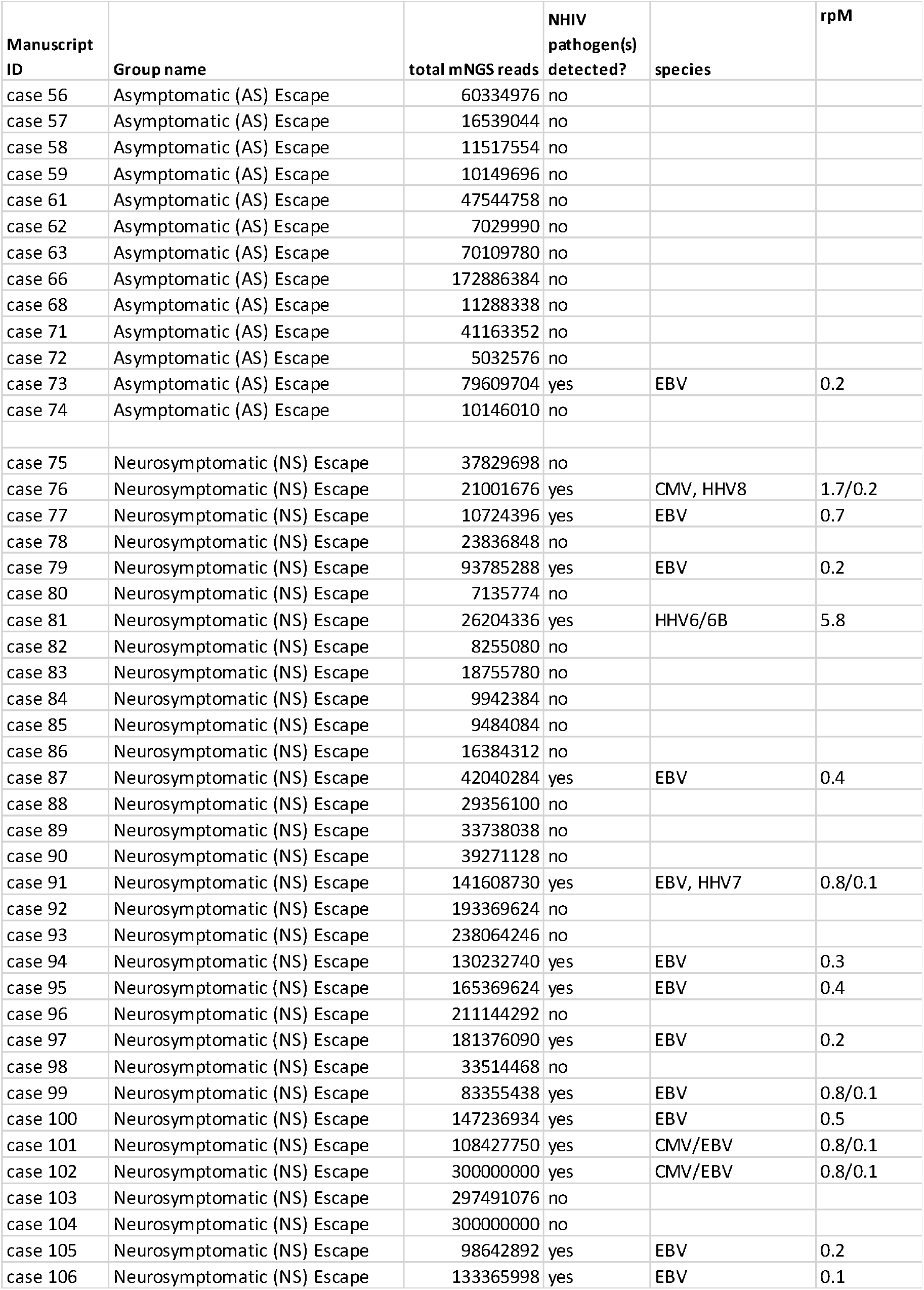
individual mNGS results for AS and NS escape subjects.

| Manuscript ID | Group name | total mNGS reads | NHIV pathogen(s) detected? | species | rpM |
| --- | --- | --- | --- | --- | --- |
| case 56 | Asymptomatic (AS) Escape | 60334976 | no |  |  |
| case 57 | Asymptomatic (AS) Escape | 16539044 | no |  |  |
| case 58 | Asymptomatic (AS) Escape | 11517554 | no |  |  |
| case 59 | Asymptomatic (AS) Escape | 10149696 | no |  |  |
| case 61 | Asymptomatic (AS) Escape | 47544758 | no |  |  |
| case 62 | Asymptomatic (AS) Escape | 7029990 | no |  |  |
| case 63 | Asymptomatic (AS) Escape | 70109780 | no |  |  |
| case 66 | Asymptomatic (AS) Escape | 172886384 | no |  |  |
| case 68 | Asymptomatic (AS) Escape | 11288338 | no |  |  |
| case 71 | Asymptomatic (AS) Escape | 41163352 | no |  |  |
| case 72 | Asymptomatic (AS) Escape | 5032576 | no |  |  |
| case 73 | Asymptomatic (AS) Escape | 79609704 | yes | EBV | 0.2 |
| case 74 | Asymptomatic (AS) Escape | 10146010 | no |  |  |
| case 75 | Neurosymptomatic (NS) Escape | 37829698 | no |  |  |
| case 76 | Neurosymptomatic (NS) Escape | 21001676 | yes | CMV, HHV8 | 1.7/0.2 |
| case 77 | Neurosymptomatic (NS) Escape | 10724396 | yes | EBV | 0.7 |
| case 78 | Neurosymptomatic (NS) Escape | 23836848 | no |  |  |
| case 79 | Neurosymptomatic (NS) Escape | 93785288 | yes | EBV | 0.2 |
| case 80 | Neurosymptomatic (NS) Escape | 7135774 | no |  |  |
| case 81 | Neurosymptomatic (NS) Escape | 26204336 | yes | HHV6/6B | 5.8 |
| case 82 | Neurosymptomatic (NS) Escape | 8255080 | no |  |  |
| case 83 | Neurosymptomatic (NS) Escape | 18755780 | no |  |  |
| case 84 | Neurosymptomatic (NS) Escape | 9942384 | no |  |  |
| case 85 | Neurosymptomatic (NS) Escape | 9484084 | no |  |  |
| case 86 | Neurosymptomatic (NS) Escape | 16384312 | no |  |  |
| case 87 | Neurosymptomatic (NS) Escape | 42040284 | yes | EBV | 0.4 |
| case 88 | Neurosymptomatic (NS) Escape | 29356100 | no |  |  |
| case 89 | Neurosymptomatic (NS) Escape | 33738038 | no |  |  |
| case 90 | Neurosymptomatic (NS) Escape | 39271128 | no |  |  |
| case 91 | Neurosymptomatic (NS) Escape | 141608730 | yes | EBV, HHV7 | 0.8/0.1 |
| case 92 | Neurosymptomatic (NS) Escape | 193369624 | no |  |  |
| case 93 | Neurosymptomatic (NS) Escape | 238064246 | no |  |  |
| case 94 | Neurosymptomatic (NS) Escape | 130232740 | yes | EBV | 0.3 |
| case 95 | Neurosymptomatic (NS) Escape | 165369624 | yes | EBV | 0.4 |
| case 96 | Neurosymptomatic (NS) Escape | 211144292 | no |  |  |
| case 97 | Neurosymptomatic (NS) Escape | 181376090 | yes | EBV | 0.2 |
| case 98 | Neurosymptomatic (NS) Escape | 33514468 | no |  |  |
| case 99 | Neurosymptomatic (NS) Escape | 83355438 | yes | EBV | 0.8/0.1 |
| case 100 | Neurosymptomatic (NS) Escape | 147236934 | yes | EBV | 0.5 |
| case 101 | Neurosymptomatic (NS) Escape | 108427750 | yes | CMV/EBV | 0.8/0.1 |
| case 102 | Neurosymptomatic (NS) Escape | 300000000 | yes | CMV/EBV | 0.8/0.1 |
| case 103 | Neurosymptomatic (NS) Escape | 297491076 | no |  |  |
| case 104 | Neurosymptomatic (NS) Escape | 300000000 | no |  |  |
| case 105 | Neurosymptomatic (NS) Escape | 98642892 | yes | EBV | 0.2 |
| case 106 | Neurosymptomatic (NS) Escape | 133365998 | yes | EBV | 0.1 |

## References

1. Valcour V, Chalermchai T, Sailasuta N, et al. Central nervous system viral invasion and inflammation during acute HIV infection. J Infect Dis. 2012;206(2):275–282. doi:10.1093/infdis/jis326

2. Ostrowski SR, Katzenstein TL, Pedersen BK, Gerstoft J, Ullum H. Residual viraemia in HIV-1-infected patients with plasma viral load ≤20 copies/ml is associated with increased blood levels of soluble immune activation markers. Scand J Immunol. 2008;68(6):652–660. doi:10.1111/j.1365-3083.2008.02184.x

3. Canestri A, Lescure FX, Jaureguiberry S, et al. Discordance between cerebral spinal fluid and plasma HIV replication in patients with neurological symptoms who are receiving suppressive antiretroviral therapy. Clin Infect Dis. 2010;50(5):773–778. doi:10.1086/650538

4. Grill MF, Price RW. Central Nervous System HIV-1 Infection. Vol 123. 1st ed. Elsevier B.V.; 2014. doi:10.1016/B978-0-444-53488-0.00023-7

5. Ferretti F, Gisslen M, Cinque P, Price RW. Cerebrospinal Fluid HIV Escape from Antiretroviral Therapy. Curr HIV/AIDS Rep. 2015;12(2):280–288. doi:10.1007/s11904-015-0267-7

6. Hagberg L, Price RW, Zetterberg H, Fuchs D, Gisslén M. Herpes zoster in HIV-1 infection: The role of CSF pleocytosis in secondary CSF escape and discordance. PLoS One. 2020;15(7):e0236162. doi:10.1371/journal.pone.0236162

7. Mukerji SS, Misra V, Lorenz D, et al. Temporal Patterns and Drug Resistance in CSF Viral Escape among ART-Experienced HIV-1 Infected Adults. J Acquir Immune Defic Syndr. 2017;75(2):246–255. doi:10.1097/QAI.0000000000001362

8. Edén A, Fuchs D, Hagberg L, et al. HIV-1 viral escape in cerebrospinal fluid of subjects on suppressive antiretroviral treatment. J Infect Dis. 2010;202(12):1819–1825. doi:10.1086/657342

9. Edén A, Nilsson S, Hagberg L, et al. Asymptomatic cerebrospinal fluid HIV-1 viral blips and viral escape during antiretroviral therapy: A longitudinal study. J Infect Dis. 2016;214(12):1822–1825. doi:10.1093/INFDIS/JIW454

10. Peluso MJ, Ferretti F, Peterson J, et al. Cerebrospinal fluid HIV escape associated with progressive neurologic dysfunction in patients on antiretroviral therapy with well controlled plasma viral load. Aids. 2012;26(14):1765–1774. doi:10.1097/QAD.0b013e328355e6b2

11. Lupia T, Milia MG, Atzori C, et al. Presence of Epstein-Barr virus DNA in cerebrospinal fluid is associated with greater HIV RNA and inflammation. Aids. 2020;34(3):373–380. doi:10.1097/QAD.0000000000002442

12. Ramachandran PS, Wilson MR. Metagenomics for neurological infections — expanding our imagination. Nat Rev Neurol. 2020;16(10):547–556. doi:10.1038/s41582-020-0374-y

13. Xu GJ, Kula T, Xu Q, et al. Comprehensive serological profiling of human populations using a synthetic human virome. 2015;348(6239). doi:10.1126/science.aaa0698

14. Schubert RD, Hawes IA, Ramachandran PS, et al. Pan-viral serology implicates enteroviruses in acute flaccid myelitis. Nat Med. 2019;25(November). doi:10.1038/s41591-019-0613-1

15. Larman HB, Zhao Z, Laserson U, et al. Autoantigen discovery with a synthetic human peptidome. Nat Biotechnol. 2011;29(6):535–541. doi:10.1038/nbt.1856

16. O’donovan B, Mandel-Brehm C, Vazquez SE, et al. High-resolution epitope mapping of anti-Hu and anti-Yo autoimmunity by programmable phage display. Brain Commun. 2020;2(2):1–16. doi:10.1093/braincomms/fcaa059

17. Gisslen M, Keating SM, Spudich S, et al. Compartmentalization of cerebrospinal fluid inflammation across the spectrum of untreated HIV-1 infection, central nervous system injury and viral suppression. PLoS One. 2021;16(5 May). doi:10.1371/journal.pone.0250987

18. Ramachandran PS, Ramesh A, Creswell F V., et al. Integrating central nervous system metagenomics and host response for diagnosis of tuberculosis meningitis and its mimics. Nat Commun. 2022;13(1):1–12. doi:10.1038/s41467-022-29353-x

19. Kalantar KL, Carvalho T, De Bourcy CFA, et al. IDseq-An open source cloud-based pipeline and analysis service for metagenomic pathogen detection and monitoring. Gigascience. 2021;9(10):1–14. doi:10.1093/GIGASCIENCE/GIAA111

20. Song E, Bartley CM, Chow RD, et al. Divergent and self-reactive immune responses in the CNS of COVID-19 patients with neurological symptoms. Cell Reports Med. 2021;2(5):100288. doi:10.1016/j.xcrm.2021.100288

21. Bodenhofer U, Bonatesta E, Horejš-Kainrath C, Hochreiter S. Msa: An R package for multiple sequence alignment. Bioinformatics. 2015;31(24):3997–3999. doi:10.1093/bioinformatics/btv494

22. Winston A, Antinori A, Cinque P, et al. Defining cerebrospinal fluid HIV RNA escape: Editorial review AIDS. Aids. 2019;33(January):S107–S111. doi:10.1097/QAD.0000000000002252

23. Hermansson L, Yilmaz A, Price RW, et al. Plasma concentration of neurofilament light chain protein decreases after switching from tenofovir disoproxil fumarate to tenofovir alafenamide fumarate. PLoS One. 2019;14(12):1–10. doi:10.1371/journal.pone.0226276

24. Peterson J, Gisslen M, Zetterberg H, et al. Cerebrospinal fluid (CSF) neuronal biomarkers across the spectrum of HIV infection: Hierarchy of injury and detection. PLoS One. 2014;9(12):1–28. doi:10.1371/journal.pone.0116081

25. Moir S, Fauci AS. B-cell responses to HIV infection. Immunol Rev. 2017;275(1):33–48. doi:10.1111/imr.12502

26. Cardozo T, Kimura T, Philpott S, Weiser B, Burger H, Zolla-Pazner S. Structural Basis for Coreceptor Selectivity by The HIV Type 1 V3 Loop. AIDS Res Hum Retroviruses. 2007;23(3):415–426. doi:10.1089/aid.2006.0130

27. Joseph SB, Kincer LP, Bowman NM, et al. Human Immunodeficiency Virus Type 1 RNA Detected in the Central Nervous System (CNS) After Years of Suppressive Antiretroviral Therapy Can Originate from a Replicating CNS Reservoir or Clonally Expanded Cells. 2018;27599(Xx):1–8. doi:10.1093/cid/ciy1066

28. Yang H, Llano A, Cedeño S, et al. Incoming HIV virion-derived Gag Spacer Peptide 2 (p1) is a target of effective CD8+ T cell antiviral responses. Cell Rep. 2021;35(6). doi:10.1016/j.celrep.2021.109103

29. Mastrangelo A, Turrini F, De Zan V, Caccia R, Gerevini S, Cinque P. Symptomatic cerebrospinal fluid escape. Aids. 2019;33(July 2018):S159–S169. doi:10.1097/QAD.0000000000002266

30. Kleines M, Scheithauer S, Schiefer J, Häusler M. Clinical application of viral cerebrospinal fluid PCR testing for diagnosis of central nervous system disorders: A retrospective 11-year experience. Diagn Microbiol Infect Dis. 2014;80(3):207–215. doi:10.1016/j.diagmicrobio.2014.07.010

31. Stam AJ, Nijhuis M, Van Den Bergh WM, Wensing AMJ. Differential genotypic evolution of HIV-1 quasispecies in cerebrospinal fluid and plasma: A systematic review. AIDS Rev. 2013;15(3):152–161.

32. Spatola M, Loos C, Cizmeci D, et al. Functional Compartmentalization of Antibodies in the Central Nervous System During Chronic HIV Infection. J Infect Dis. 2022;226(4):738–750. doi:10.1093/infdis/jiac138

33. Joseph SB, Arrildt KT, Sturdevant CB, Swanstrom R. HIV-1 target cells in the CNS. J Neurovirol. 2015;21(3):276–289. doi:10.1007/s13365-014-0287-x

34. Gorry PR, Taylor J, Holm GH, et al. Increased CCR5 Affinity and Reduced CCR5/CD4 Dependence of a Neurovirulent Primary Human Immunodeficiency Virus Type 1 Isolate. J Virol. 2002;76(12):6277–6292. doi:10.1128/jvi.76.12.6277-6292.2002

35. Mzoughi O, Teixido M, Planès R, et al. Trimeric heptad repeat synthetic peptides HR1 and HR2 efficiently inhibit HIV-1 entry. Biosci Rep. 2019;39(9):1–15. doi:10.1042/BSR20192196/220430

36. Eshleman SH, Laeyendecker O, Kammers K, et al. Comprehensive Profiling of HIV Antibody Evolution Article Comprehensive Profiling of HIV Antibody Evolution. CellReports. 2019;27(5):1422-1433.e4. doi:10.1016/j.celrep.2019.03.097

37. Dobyns WB, Aldinger KA, Ishak GE, et al. MACF1 Mutations Encoding Highly Conserved Zinc-Binding Residues of the GAR Domain Cause Defects in Neuronal Migration and Axon Guidance. Am J Hum Genet. 2018;103(6):1009–1021. doi:10.1016/j.ajhg.2018.10.019

